# Burden of COVID-19 and Case Fatality Rate in Pune India: An Analysis of First and Second Wave of the Pandemic

**DOI:** 10.1101/2021.06.21.21259225

**Authors:** Prasad Bogam, Aparna Joshi, Sanket Nagarkar, Divyashri Jain, Nikhil Gupte, LS Shashidhara, Joy Merwin Monteiro, Vidya Mave

## Abstract

**Background:** The recent second wave in India in April-May 2021 placed an unprecedented burden on the Indian health systems. However, limited data exist on the epidemiology of the COVID-19 pandemic from the first wave through the second wave in India. With detailed epidemiologic data, we aimed to assess trends in incident cases and case fatality, its risk between pandemic waves in Pune, an epicenter of COVID-19 cases in India, a country with the second-largest absolute burden worldwide.

**Methods:** Programmatic COVID-19 data from Pune city between the first wave (March 09^th^ 2020-October 31^st^, 2020), maintenance phase (November 01^st^ 2020-February 14^th^, 2021), the second wave (February 15^th^, 2021-May 31^st^, 2021) were assessed for trends of incident cases, time-to-death, and case fatality rate (CFR). In addition, Poisson regression models adjusted for age and gender were used to determine the independent effect of pandemic waves on mortality.

**Results:** Of 465,192 COVID-19 cases, 162,182 (35%) were reported in the first wave, and 4,146 (2.5%) died among them; Maintenance period registered 27,517 (6%) cases with 590 (2.1%) deaths; Second wave reported 275,493 (59%) cases and 3184 (1.1%) deaths (p<0.01). The overall CFR was 1.16 per 1000 person-days (PD), which declined from 1.80 per 1000 PD during the first wave to 0.77 per 1000 PD in the second wave. The risk of death was 1.49 times higher during the first wave (adjusted case fatality rate ratio-aCFRR,1.49; 95% CI: 1.37–1.62) and 35% lower in the second wave (aCFRR, 0.65; 95% CI: 0.59 – 0.70), compared to the maintenance phase.

**Interpretation:** The absolute burden of COVID-19 cases and deaths were more significant in the second wave in Pune, India; however, the CFR declined as the pandemic progressed. Nevertheless, investigating newer therapies and implementing mass vaccinations against COVID-19 are urgently needed.

## Introduction

The global COVID-19 cases exceeded 171 million as of May 31^st^, 2021, leading to a loss of more than 3.69 million human lives.^1^ Data to date suggests that elderly males and those with comorbidities are at higher risk for developing poor clinical outcomes, including mortality, during and after acute SARS-CoV-2 infection.^2^ India shares about 16% of the reported global burden of COVID-19 cases and ranks second in the absolute number of cumulative COVID-19 cases. The first wave of the COVID-19 pandemic commenced with increased detection of cases in January-March 2020, and after the September 2020 peak, cases declined by the end of October 2020.^3^ We previously reported that the national lockdown in India delayed the peak of the first wave of the pandemic by approximately eight weeks.^4^ Subsequently, a low COVID-19 incidence period occurred between November and mid-February. However, the recent second wave in India in April-May 2021 placed an unprecedented burden on the Indian health systems, which contributed almost 47% of single-day incident cases in the world during its peak.^5^ Limited data exist on the epidemiology of the progression of the COVID-19 pandemic from the first wave through the second wave in India.

With 334,608 COVID-19 related deaths to date, India ranks third in the absolute number of deaths.^1^ Interestingly, the observed case fatality ratio at 1.2% is the lowest among the top 20 high burden countries.^6^ In general, the lower COVID-19 case fatality ratio reported in India can be attributed to the younger demographic profile of the population compared to that of other high-income countries.^7^ However, the explanation might be far more complex given the data collecting and reporting structure, prevalence of comorbid diseases in India, testing strategy, sex differences, and death attribution to COVID-19 infections.^8-9^ While emerging evidence from western countries supported the use of glucocorticoids, antivirals, monoclonal antibodies, and optimal critical care illness management of COVID-19,^10-12^, limited data exists on its impact in resource-limited countries at a programmatic level. As Pune city in India has a system of reporting detailed mortality data, we sought to describe an epidemiological analysis of the burden and COVID-19 mortality data comparing the first and second wave in the city, one of India’s epicenters of COVID-19 cases.

## Methods

Between March 01^st^, 2020, and May 31^st^, 2021, we conducted a retrospective analysis of the COVID-19 mortality data from Pune city, India. As part of the COVID-19 surveillance program, the public and private hospitals under the jurisdiction of Pune Municipal Corporation (PMC) in Pune city, India, shared daily reports of COVID-19 incident cases and related deaths with PMC. The data elements included age, sex, date of diagnosis, and for those who died, date of death. Aggregate daily data on the number of SARS-CoV-2 tests, new diagnoses, active cases, and critical care cases were compiled at the city level. The individual hospital-level data on deaths were collected centrally, and the cause of death remark was curated to elicit the comorbidities from the remarks.

The details of programmatic management of the pandemic in India, including lockdowns during the early phases, have been described elsewhere.^4,13^ In brief, between 25-March-2020 and 31-May-2020, the nationwide lockdown was implemented, and the regional lockdown of two weeks was implemented in the Pune region in July 2020. During the second wave of the pandemic, regional lockdown in Maharashtra state, including Pune, was implemented in April-May 2021. In the early phases of the first wave, COVID-19 testing was available only for symptomatic close contacts of laboratory-confirmed cases and international travelers with a history of travel to COVID-19 affected countries who develop respiratory symptoms.^14^ However, in the last week of March 2020, the testing expanded to symptomatic healthcare workers, all hospitalized patients with severe acute respiratory illness, and the asymptomatic direct and high-risk contacts of confirmed cases.^15^ Subsequently, the Indian Council of Medical Research (ICMR) devised a strategy to increase accessibility and availability of testing by engaging with Government and private laboratories to add the optimum number of testing centers in the country.^16^ Rapid antigen testing was added into the testing strategy in April 2020 to test the symptomatic in hotspot areas.^17^ In the later months of the pandemic, rapid antigen tests and RT-PCR were suggested to test all individuals residing in the containment zones as part of community surveillance. During the second wave, RT-PCR was advised for all symptomatic testing negative on rapid antigen test.

Home isolation for mild/pre-symptomatic patients was permitted starting 27-April-2020; hospitalization was only recommended to those with moderate and severe COVID-19 illness. The moderate disease was defined as the presence of breathlessness, respiratory rate (RR) of ≥ 24/ min, or oxygen saturation (SpO2) of <93% on room air; severe disease was defined as the presence of breathlessness, RR of > 30/ min or SpO2 of <90% on room air. The local authorities compiled the mandated data on testing and death outcomes among cases from individual facility level-testing centers, laboratories, hospitals, care centers and shared nationally.

### COVID-19 management

The Ministry of Health and Family Welfare (MoHFW) issued guidelines on clinical management of COVID-19 starting 17-Mar-2020 and subsequently revised them based on the available evidence.^18^ Initially, the treatment strategies were primarily based on early supportive therapy with supplemental oxygen, empirical antimicrobials, and management of acute respiratory distress syndrome (ARDS) with non-invasive mechanical ventilation or invasive mechanical ventilation if required. Glucocorticoids were also recommended for a short period (3-5 days) in deteriorating patients. Hydroxychloroquine (HCQ) was repurposed and indicated as an off-label treatment for COVID combined with Azithromycin among patients with severe illness requiring ICU admissions. In the subsequent months, the treatment strategies evolved, and the indicated use of HCQ shifted from severe cases to milder cases or at-risk populations such as healthcare workers. On June 13th and 27th, MoHFW revised guidelines and recommended a 0.1-0.2mg/kg dose of dexamethasone or its equivalent glucocorticoid among patients with increasing oxygen requirements. In addition, multiple investigational therapies were advised as ad-hoc, including remdesivir (an antiviral agent), convalescent plasma therapy, and tocilizumab (an anti-interleukin-6 monoclonal antibody) as off label treatment for COVID-19 for moderate disease.

### Statistical analysis

For this analysis, we divided the COVID-19 pandemic into three phases based on the trend of incident cases and test positivity rates in Pune: the first phase representing the first wave of the pandemic from March 09^th^,2020, until October 31^st^, 2020, followed by the maintenance phase from November 01^st^,2020 until February 15^th^, 2021. The last phase starting February 16^th^,2021, represents the second wave currently ongoing in the country, and this analysis was censored for May 31^st^, 2021, for incident cases (**Supplementary Table 1**). Characteristics of COVID-19 cases in the first wave, maintenance phase, and second-wave were summarized using frequencies and compared with a Chi-Squared test. The overall case burden for India abstracted from publicly available data was plotted over the study period. The test positivity rates were derived and plotted from the aggregated daily public reports. Case fatality ratio was measured by dividing the total deaths from total diagnosed cases. The ratio of critical care cases to daily active cases was plotted. The COVID-19 deaths were censored for June 07^th^, 2021. Case fatality rate (CFR) overall and age groups, sex, and the three pandemic phases were calculated as the total number of deaths divided by total cases diagnosed per 1000 person-days (PD). Cumulative time to death was estimated using the Kaplan-Meier product-limit estimator and compared within the risk groups using a log-rank test. Univariable and multivariable Poisson regression was used to assess the effects of sex, age categories, and three pandemic phases on mortality for the overall pandemic and stratified by three phases of the pandemic. Furthermore, we compared the CFR before and after the national guidelines for intravenous steroids and their impact on mortality using logistic regression analysis. The Indian Institute of Science Education and Research ethics committee, Pune, India, approved this analysis of the public health monitoring program.

## Results

### COVID-19 in the first wave, second wave, and maintenance phase

During the study period, 465,192 COVID-19 cases were diagnosed in Pune city; 162,182 (35% of total cases) in the first wave, 27,517 (6%) in the maintenance period, and 275,493 (59%) in the second wave. The trends in COVID-19 case detection in Pune city and that of India are shown in **Figure 1a**, and the peak of the second wave was delayed by 12 days for India compared to Pune city. Overall, the test positivity of cases was 19%; it was highest at 22% (p<0.01) during the first wave, followed by the 19% (p<0.01) in the second wave and 8% (p=0.18) in the maintenance period (**Supplementary Figure 1**). As shown in **Table 1**, the 30-45 years age group (p<0.01) and males (p<0.01) contributed to most COVID-19 cases across all three phases of the pandemic. The most increase in the proportion of weekly cases was observed among the 30-44 years age group (p<0.01) followed by 45-64 (p=0.02); 65-85 (p=0.003) year age group during the analysis period **(Figure 1b)**. The ratio of critical care cases to the active cases (**Figure 1c**) was 6% in the first wave, 8% in the maintenance phase, and 3% in the second wave (p<0.01).

**Table 1 -.** Characteristics of COVID-19 incident cases and deaths in the first wave, maintenance phase, and second wave in Pune, India.

| Characteristics | Total Cases |  | First Wave |  | Maintenance phase |  | Second Wave |  |
| --- | --- | --- | --- | --- | --- | --- | --- | --- |
|  | Cases n (%) | Deaths n (%) | Cases n (%) | Deaths n (%) | Cases n (%) | Deaths n (%) | Cases n (%) | Deaths n (%) |
| Overall | 465192 | 7920 (2) | 162182 (35) | 4146 (3) | 27517 (6) | 590 (2) | 275493 (59) | 3184 (1) |
| Median Age, IQR | 38 (28 – 52) | 65 (54 – 73) | 38 (27 – 52) | 65 (55 – 73) | 41 (30 – 55) | 66 (55 – 75) | 38 (28 – 52) | 64 (52 – 73) |
| Age categories |  |  |  |  |  |  |  |  |
| 0 – 17 | 43859 (9) | 37 (0) | 17577 (11) | 28 (0) | 2105 (8) | 1 (0) | 24177 (9) | 8 (0) |
| 18 – 29 | 92218 (20) | 82 (0) | 31304 (19) | 29 (0) | 4708 (17) | 14 (0) | 56206 (20) | 39 (0) |
| 30 – 44 | 155588 (33) | 726 (1) | 51228 (32) | 306 (1) | 8601 (31) | 51 (1) | 95759 (35) | 369 (0) |
| 45 – 64 | 123015 (26) | 3015 (3) | 45115 (28) | 1618 (4) | 8594 (31) | 196 (2) | 69306 (25) | 1201 (2) |
| 65 – 85 | 47991 (10) | 3755 (8) | 16287 (10) | 2026 (12) | 3359 (12) | 300 (9) | 28345 (10) | 1429 (5) |
| >85 | 2521 (1) | 305 (12) | 671 (0) | 139 (21) | 150 (1) | 28 (19) | 1700 (1) | 138 (8) |
| Gender |  |  |  |  |  |  |  |  |
| Female | 199446 (43) | 2729 (1) | 68089 (42) | 1342 (2) | 11661 (42) | 201 (2) | 119696 (43) | 1186 (1) |
| Male | 265623 (57) | 5185 (2) | 94071 (58) | 2799 (3) | 15855 (58) | 389 (3) | 155697 (57) | 1997 (1) |

**Figure 1a -.**
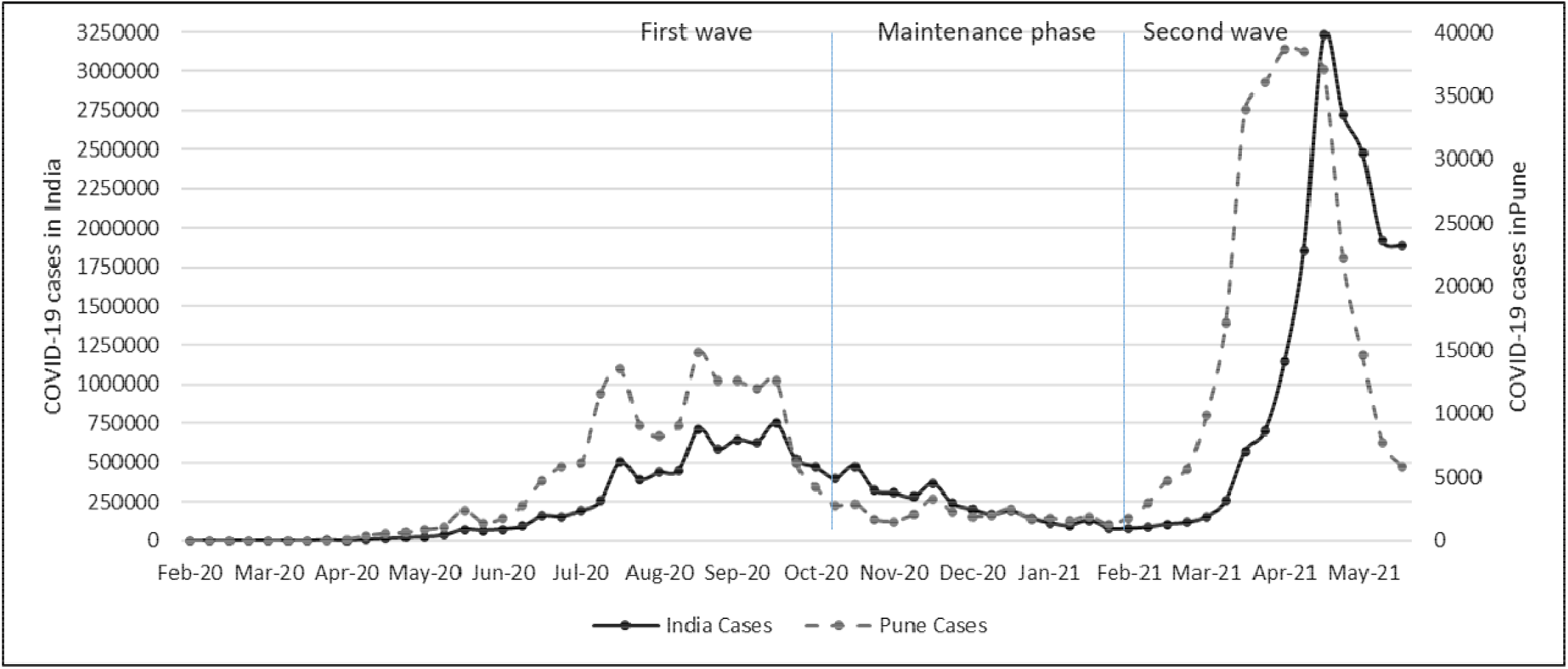
Weekly incident COVID-19 cases in Pune and India from March 09^th^,2020, until May 31^st^,2021. The first wave of the pandemic is between March 09^th^,2020, and October 31^st^, 2020; the maintenance phase is between November 01^st^,2020, and February 15^th^, 2021. The second wave is between February 16^th^,2021, and May 31^st^, 2021, and this analysis was censored for May 1^st^, 2021, for incident cases and June 07^th^, 2021, for deaths.

**Figure 1b –.**
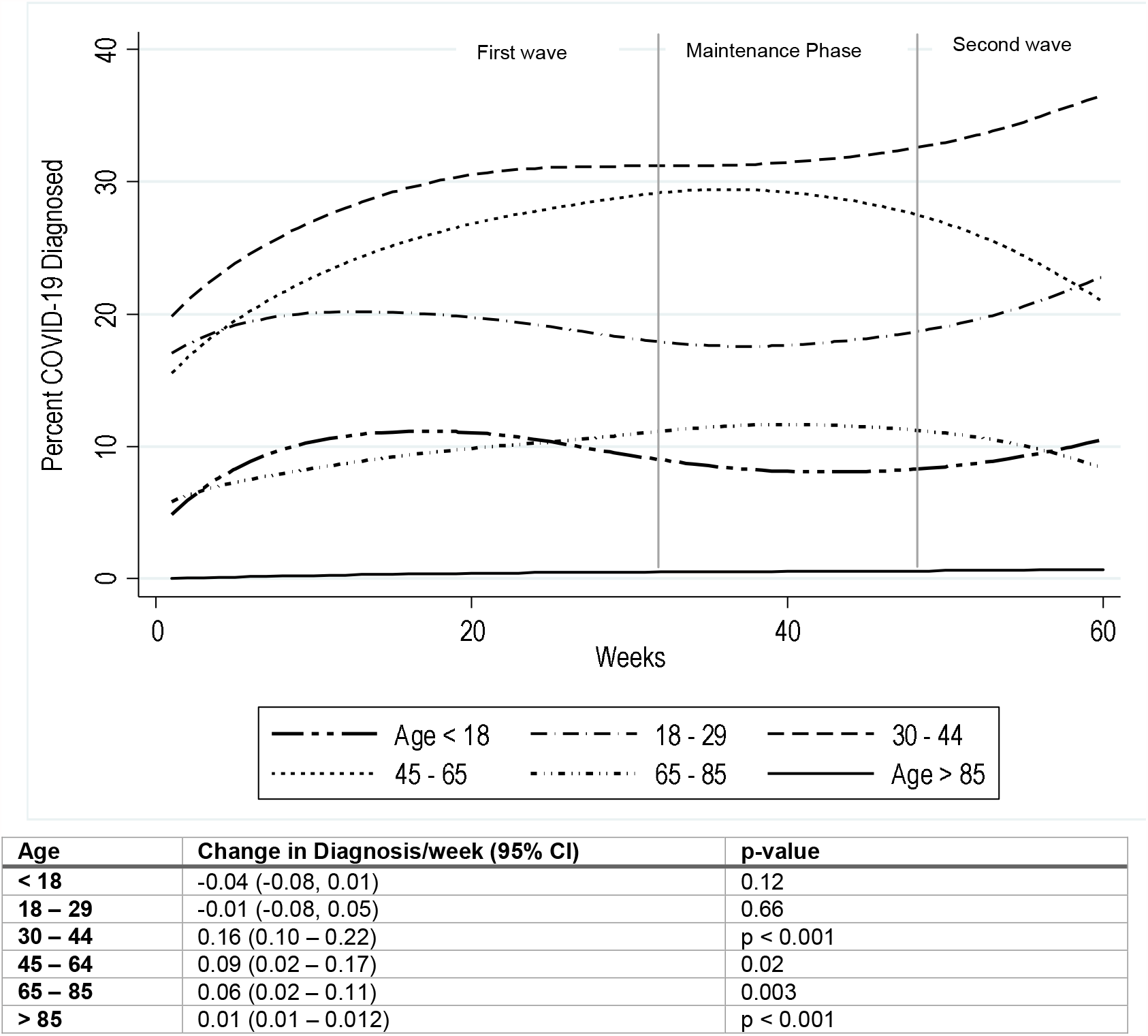
Weekly incidents of COVID-19 cases in Pune among age groups from March 09^th^,2020 until May 31^st^,2021 The first wave of the pandemic is between March 09^th^,2020, and October 31^st^, 2020; the maintenance phase is between November 01^st^,2020, and February 15^th^, 2021. The second wave is between February 16^th^,2021, and May 31^st^, 2021, and this analysis was censored for May 1^st^, 2021, for incident cases and June 07^th^, 2021, for deaths.

**Figure 1c:**
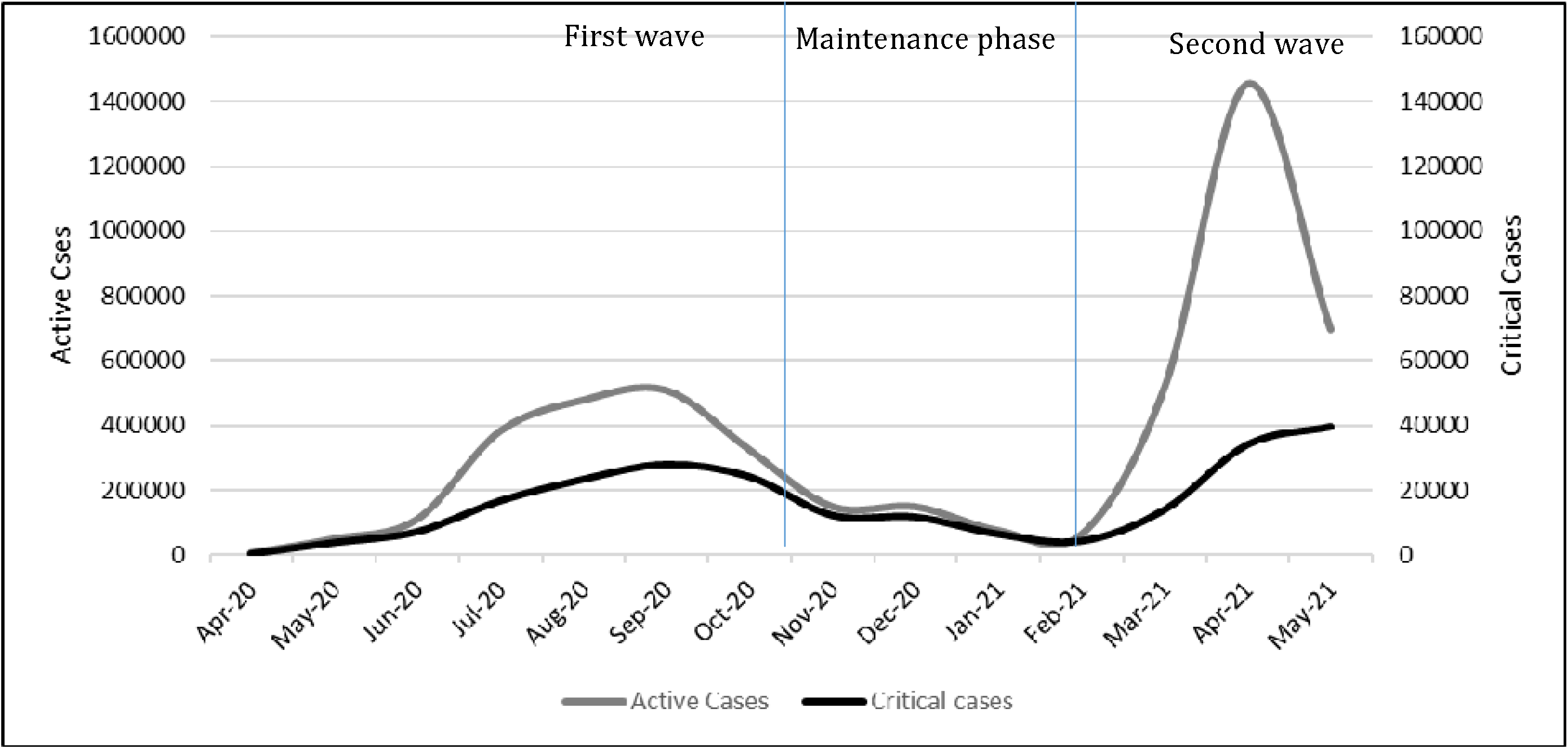
Monthly Active cases and Critical Care cases in Pune City. The first wave of the pandemic is between March 09^th^,2020, and October 31^st^, 2020; the maintenance phase is between November 01^st^,2020, and February 15^th^, 2021. The second wave is between February 16^th^,2021, and May 31^st^, 2021, and this analysis was censored for May 1^st^, 2021, for incident cases and June 07^th^, 2021, for deaths.

### Death characteristics and time to death

Of 465,192 cases,7,920 (1.7%) died. The median age of those who died was 65 (IQR, 54 - 73) years; with almost half of the deaths, 3,755 (47%) reported between 65-85 years of age, and 5185 (65%) were males. Of these, 3,704 (47%) had one or more comorbidities; 2,699 (34%) had hypertension and 2,239 (28%) had diabetes **(Supplementary Figure 2)**. Among those who died, the median time to death from diagnosis was six days (IQR, 2-11). As shown in **Figure 2a**, time to death was slightly different for males vs. females (6 (IQR, 2-12) vs. 5 (IQR, 2-11) days, p<.01). For age group categories, median time to deaths in days was in the following order: 6 (IQR, 3-10) for >85 years; 6 (IQR, 2-11) for 65-85 years; 6(2-12) for 45-64 years; 6 (1-12) for 30-44 years; 3 (0-9) for 18-29 years; 3 (0-7) for <17 years (**Figure 2b**, p<0.01).

**Figure 2a.**
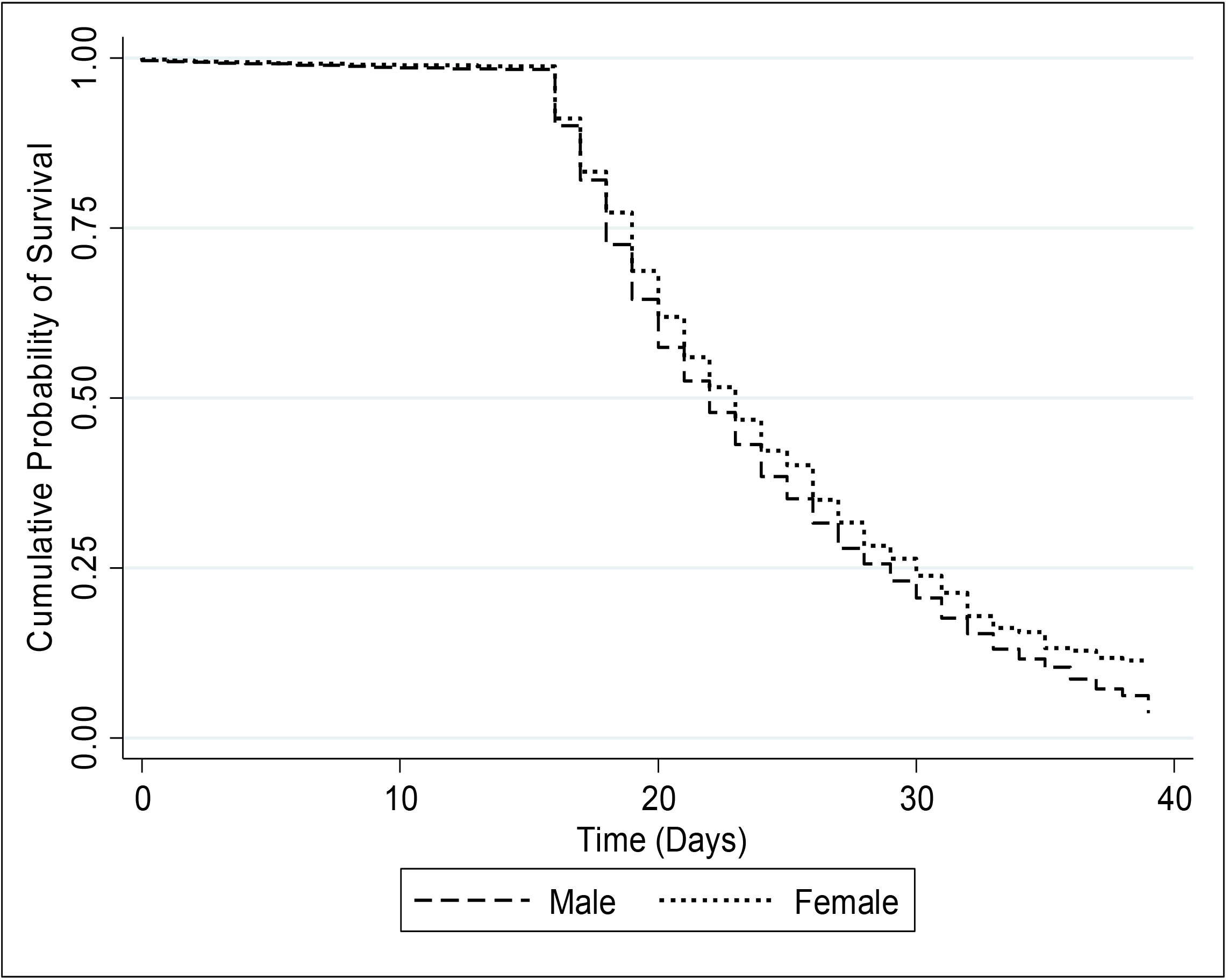
Kaplan Meier curve showing median time to death from diagnosis among males vs females.

**Figure 2b.**
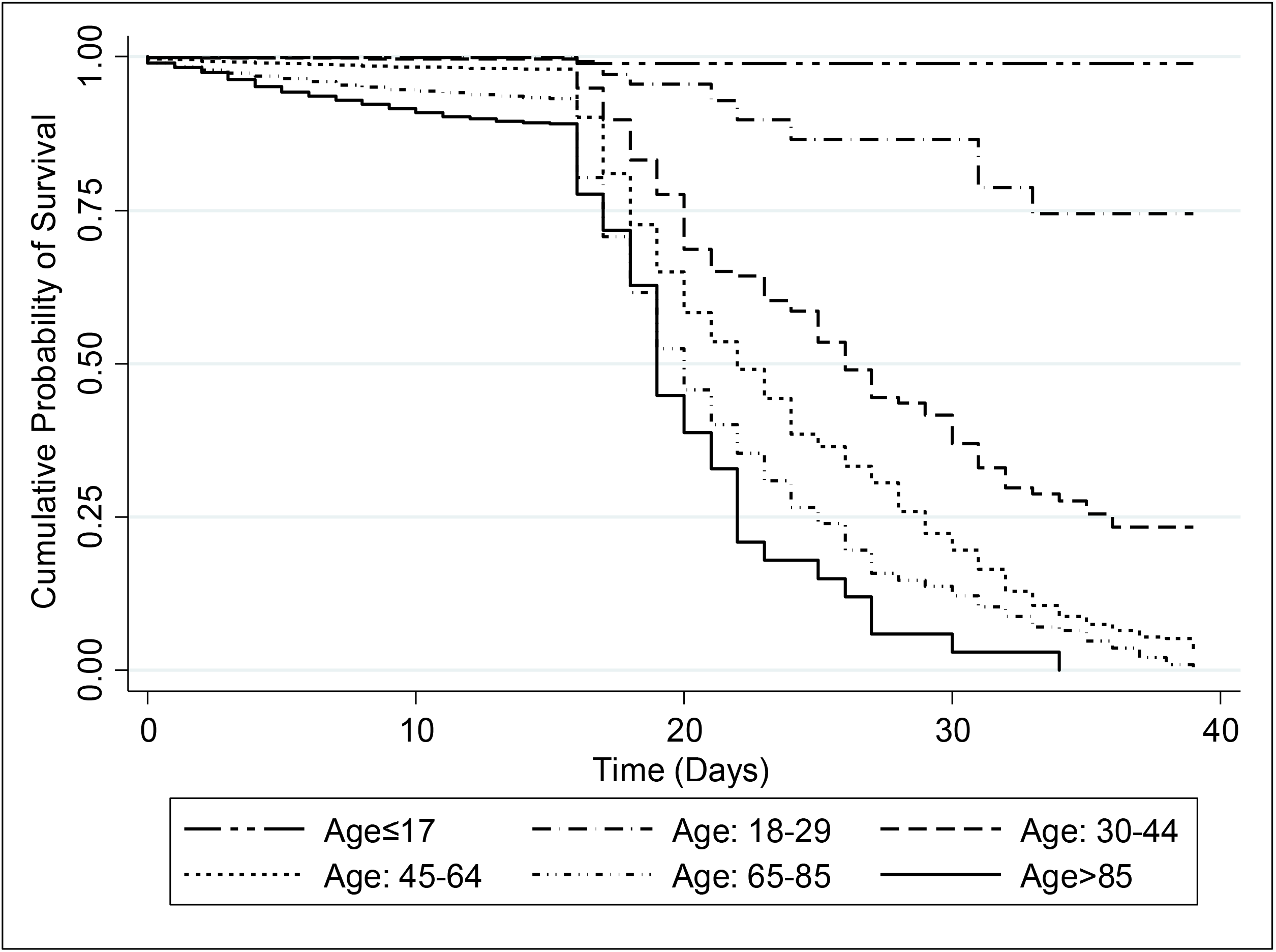
Kaplan Meier curve showing median time to death from diagnosis among different age categories.

### Case Fatality Rate and Risk Factors

Of 7,920 who died, 4,146 (52%) died in the first wave, 590 (8%) in the maintenance phase and 3,184 (40%) in the second wave (p<0.01). The case fatality rate rapidly declined from 3.91 per 1000 PD (95% CI; 3.56 - 4.25) to 1.66 per 1000 PD (95% CI: 1.61 - 1.72) during the first wave after the introduction of clinical management focused mainly on intravenous steroids use for severe COVID-19 in June 2020 (**Figure 3a**, p<0.01). It further declined to 0.77 per 1000 PD in the second wave. Moreover, the case fatality rate was 3.53 times greater before intravenous steroid use for severe illness (CFRR, 3.53, 95% CI 3.23-3.86).

**Figure 3a.**
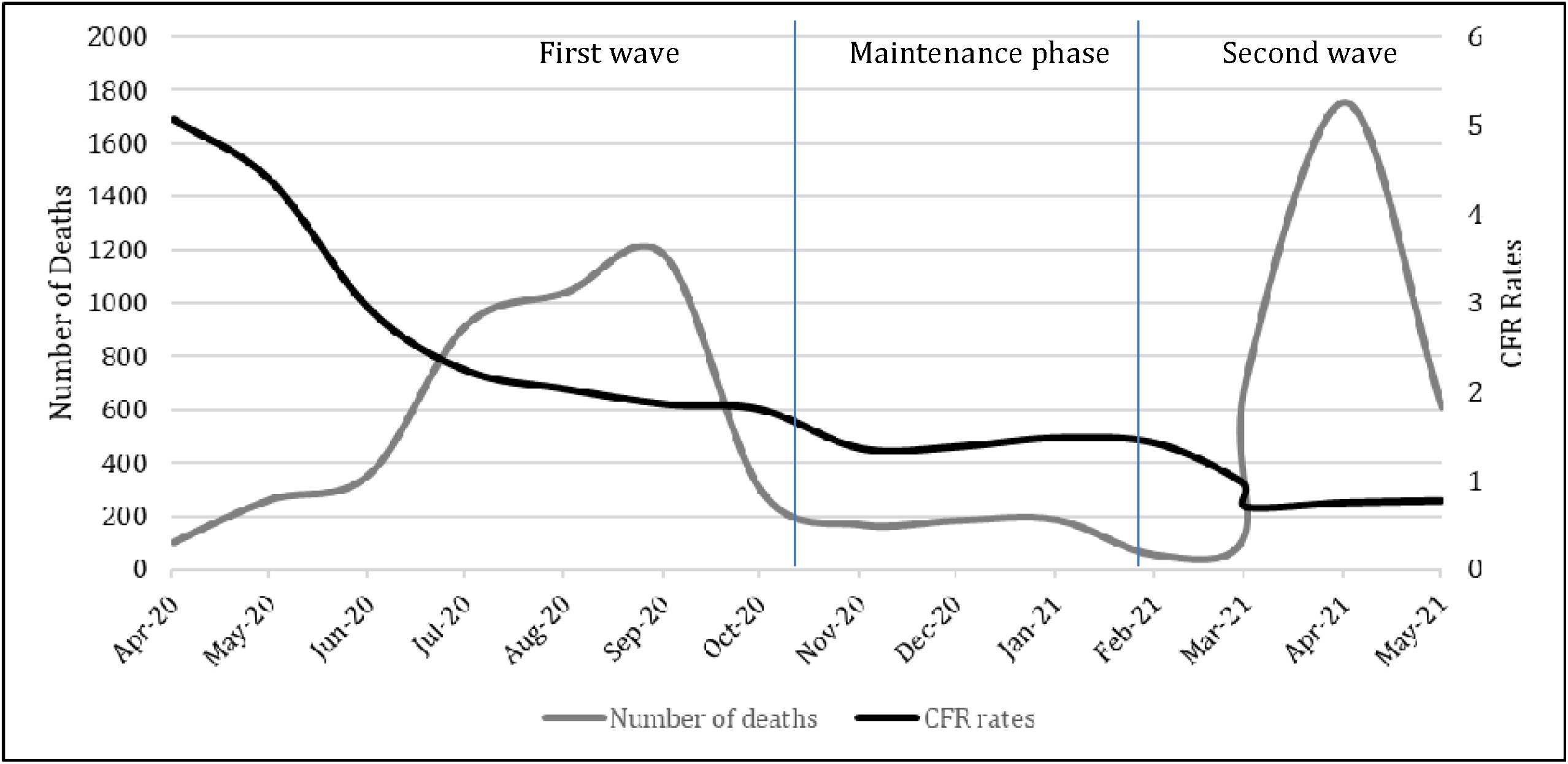
Cumulative Case Fatality Rate (per 1000 person-days) and monthly deaths in Pune. The first wave of the pandemic is between March 09^th^,2020, and October 31^st^, 2020; the maintenance phase is between November 01^st^,2020, and February 15^th^, 2021. The second wave is between February 16^th^,2021, and May 31^st^, 2021, and this analysis was censored for May 31^st^, 2021, for incident cases and June 07^th^, 2021, for deaths.

Overall case fatality rate per 1000 person-days (PD) was 1.16; it was lowest for 0-17 & 18-39 age groups at 0.06/ 1000 PD and highest for ages >85 years at 8.62/ 1000 PD (**Table 2**). The rate decreased over time; 1.80/ 1000 PD in the first wave, 1.42/ 1000 PD in the maintenance phase, and 0.77/ 1000 PD in the second wave. Age group 65-85 saw the highest decline in CFR over time (**Figure 3b**). In Poisson regression models, males (adjusted case fatality rate ratio [(aCFRR], 1.46; 95% CI: 1.40 – 1.53); ages 65-85 years (aCFRR, 90.15; 95% CI: 72.44 – 112.20) and age >85 years (aCFRR, 155.04; 95% CI: 121.49 - 197.86) had increased risk of mortality (**Table 2**). Furthermore, the risk for deaths was 1.49 times higher for the first wave (aCFRR,1.49; 95% CI: 1.37 – 1.62) as compared to the maintenance phase, while it was almost 35% lower for the second wave (aCFRR, 0.65; 95% CI:0.59 – 0.70) (**Table 2**). Furthermore, the fatality rate is 47% lower in the second wave than in the first wave (aCFRR, 0.43; CI: 0.41-0.45). Risk factor analysis for deaths by three analysis periods did not change significantly except for the 0-17 years age group (aCFRR, 1.7; 95% CI: 1.0 – 2.9) who had a higher likelihood of deaths in the first wave compared to the maintenance phase (**Supplementary Table 2**).

**Table 2-.** Factors Associated with COVID-19 Mortality using Poisson Regression Analysis in Pune city, India.

| Characteristics | Time to Death<br>Median (IQR) | CFR/1000 Person<br>Days (95% CI) | Unadjusted case fatality rate<br>ratio (95% CI) | Adjusted case fatality rate<br>ratio (95% CI) |
| --- | --- | --- | --- | --- |
| Overall | 6 (2 – 11) | 1.16 (1.13 – 1.19) | - |  |
| Age, median (IQR) |  | - | 1.073 (1.072 – 1.074) | - |
| Age categories |  |  |  |  |
| 0 – 17 | 3 (0 – 7) | 0.06 (0.04 – 0.08) | 0.95 (0.65 – 1.41) | 0.92 (0.62 – 1.35) |
| 18 – 29 | 3 (0 – 9) | 0.06 (0.05 – 0.07) | 1 | 1 |
| 30 – 44 | 6 (1 – 12) | 0.32 (0.29 – 0.34) | 5.24 (4.17 – 6.58) | 5.23 (4.16 – 6.57) |
| 45 – 64 | 6 (2 – 12) | 1.67 (1.61 – 1.73) | 27.72 (22.26 – 34.51) | 26.96 (21.65 – 33.58) |
| 65 – 85 | 6 (2 – 11) | 5.45 (5.28 – 5.63) | 90.38 (72.62 – 112.48) | 90.15 (72.44 – 112.20) |
| >85 | 6 (3 – 10) | 8.62 (7.68 – 9.64) | 142.85 (111.94 – 182.29) | 155.04 (121.49 – 197.86) |
| Gender |  |  |  |  |
| Female | 5 (2 – 11) | 0.93 (0.89 – 0.97) | 1 | 1 |
| Male | 6 (2 – 12) | 1.33 (1.29 – 1.37) | 1.43 (1.36 – 1.50) | 1.46 (1.40 – 1.53) |

| Period |  |  |  |  |
| --- | --- | --- | --- | --- |
| First wave | 6 (2 – 11) | 1.80 (1.75 - 1.86) | 1.27 (1.16 – 1.38) | 1.49 (1.37 – 1.62) |
| Maintenance | 7 (2 – 13) | 1.42 (1.31 - 1.54) | 1 | 1 |
| Second wave | 6 (2 – 11) | 0.77 (0.75 - 0.80) | 0.54 (0.50 – 0.59) | 0.65 (0.59 – 0.70) |

**Figure 3b.**
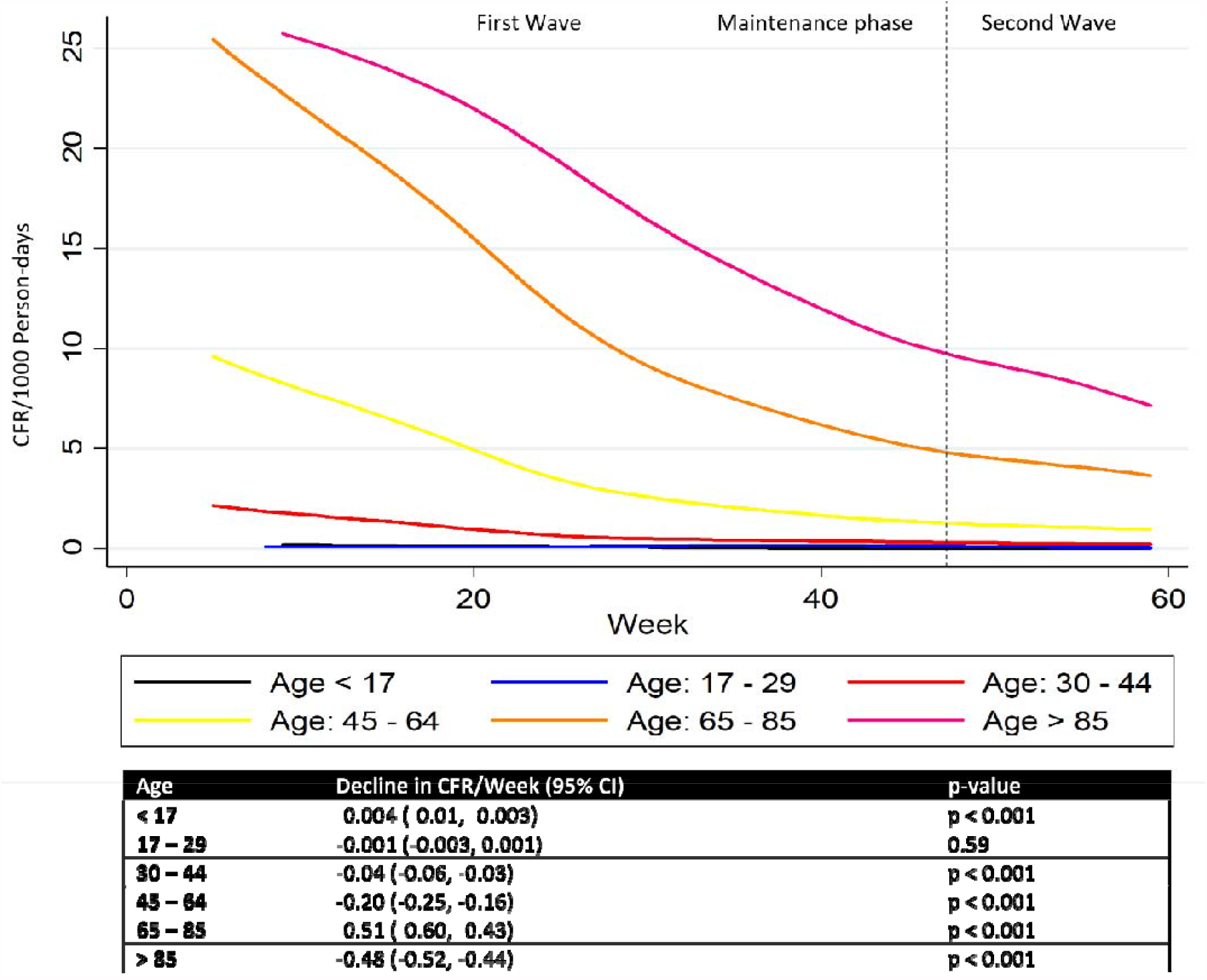
Case Fatality Rate (per 1000 person-days) over time among different age categories. The first wave of the pandemic is between March 09^th^,2020, and October 31^st^, 2020; the maintenance phase is between November 01^st^,2020, and February 15^th^, 2021. The second wave is between February 16^th^,2021, and May 31^st^, 2021, and this analysis was censored for May 31^st^, 2021, for incident cases and June 07^th^, 2021, for deaths.

## Discussion

Our report encompasses the full spectrum of Pune city’s COVID-19 pandemic up to close to the end of the second wave. The trend of COVID-19 incidence cases in Pune city followed a pattern like that of India with two exceptions. First, the peak of the second wave in Pune occurred about 12 days earlier than the country. Second, there was a slight dip in cases during the first wave corresponding to Pune regional lockdown.^4^ The first wave of the COVID-19 pandemic lasted 7-8 months, and the maintenance phase had low COVID-19 incidence. The second wave had exponential growth of cases and has the world’s largest peak and the absolute number of cases.^1^ The overall case fatality ratio was 1.7% in Pune city, higher than the reported nationwide case fatality ratio of 1.2%. Notably, the CFR declined sharply during the first wave, but it was 1.49 times higher than the phase between two pandemic waves. Furthermore, the CFR of the second wave was 35% lower than the phase between the pandemic waves and 61% lower than the first wave.

The reasons for the decline in CFR as the pandemic progressed are unclear and need further exploration. First, the proportion of COVID-19 cases among middle-aged adults (30-44 years) whose risk of deaths is relatively lower increased over time, resulting in fewer deaths. Further, the ratio of critical care cases to total active cases during the second wave was lower compared to the first wave. Second, since the first wave ended, the predominant circulating strains in India, a new variant of SARS-CoV-2-B.1.617, are known to be highly transmissible, as demonstrated by the rapid growth of cases during the second wave. Although the World Health Organization has recently categorized this strain as a variant of concern, the severity of the disease associated with this strain is yet to be investigated.^19-21^ But, our data suggest that the COVID-19 caused by SARS-CoV-2-B.1.617 may be less severe and hence less fatal. Third, during the pandemic, our test positivity analysis indicates that more cases were identified during the second wave with a lower test positivity rate than the first wave. Hence the CFR is likely to be closer to the infection fatality rate in the second wave,^22^ indicating that the increased availability of testing may have led to early diagnosis and access to care preventing further complications and deaths.

The sharp decline of COVID-19 mortality during the first wave also appears to coincide with the availability of more information and revised national guidelines on clinical management such as optimal use of corticosteroid, antivirals, and appropriate early supportive treatment following hospitalization of moderate and severe COVID-19.^23, 24,25^ Indeed, before the nationwide recommendation of advanced clinical management in early June 2020, the case fatality rate was 3.53 times higher during the first wave, before becoming stable at ∼2%.

The relatively higher cumulative case-fatality ratio observed in Pune city at 1.7% than the countrywide reported ratio of ∼1% can be explained by access to better curated and granular data at the Pune city level. Although it was much lower than western counterparts, consistent with prior studies,^26-27,^ the oldest age groups, particularly those above 65, had the highest likelihood of deaths across the pandemic and the most decline in the CFR over time.^28^ Sex bias in a higher likelihood of mortality among males was also noted, similar to prior studies,^29^ although time to death was longer for both older age groups and males.^13^ Moreover, the risk of death among children was lowest overall.^30^ Notably, the children had a 1.75 times risk of mortality during the first wave compared to young adults; however, this needs to be interpreted with caution as the proportion of children with COVID-19 were comparatively smaller, and comorbid conditions for them were not captured.

Even though the older adults had the highest risk of deaths, the proportion of deaths among the incident cases in the 18-65 years age group and males who are traditionally the wage-earners of the Indian family were closer to 50% and 66% respectively. Furthermore, considering the emerging reports of post COVID-19 sequelae and associated morbidity and mortality for this group,^31^ the economic repercussions of the pandemic may remain for the foreseeable future in India.^32^ Notably, the total disease burden and proportional critical cases were significantly higher during the second wave that required hospital beds, oxygenation, ventilator support, and health care workers to manage the illness. In this context, our report gives an insight into how the Indian health system had to manage the unprecedented burden placed by the second wave of COVID-19 pandemic widely publicized in Indian and global media reports.^33-34.^

Strengths of our analysis include the availability of more complete data enabling comparative analysis of first and second waves with that of the maintenance phase of the pandemic. While the period demarcation for these waves can be countered as arbitrary, arbitrary delineation of pandemic waves has been done worldwide to assess the progression of the pandemic.^35^ To reduce period bias, we used trends in weekly case data and test positivity to assign three phases for this analysis. The absence of data on comorbidities among COVID-19 survivors limited our ability to assess the relationship between comorbidities and mortality. However, we show that over half had one or more comorbidities among those who died, suggesting the relationship between comorbidities and mortality.^36^ Only aggregate data of critical cases were available; detailed data on hospitalization, oxygenation, and ventilator support requirements were unavailable. Furthermore, data were unavailable on SARS-CoV-2 variants and their impact on case burden and deaths and whether providers uniformly implemented the MOHFW recommendations in Pune city. The majority of deaths (99%) occurred in the hospital settings in this dataset; thus, COVID-19 deaths in the community may not have been captured, thus underreporting of COVID-19 deaths for Pune city cannot be ruled out. Despite these limitations, to our knowledge, this is the first real-world epidemiologic analysis of the progression of the pandemic and related mortality in a COVID-19 hotspot city, one of many such affected cities in India.

In conclusion, we demonstrate that as the burden of COVID-19 expanded from the first wave to the second wave, the CFR declined. Our report suggests the role of evolving therapeutics in the reduction of CFT, but further confirmation is needed. In addition, investigation of the impact of predominant circulating strains on mortality is urgently required. Notably, our report highlights the need for healthcare preparedness, including hospital beds, oxygenation, ventilator support, and health care workers to manage the pandemic of this scale worldwide. Further, there is an urgent need to investigate newer or repurposed therapies and implement mass vaccinations to halt the morbidity and mortality of the devastating COVID-19 pandemic in India and worldwide.

## Data Availability

Data will be available upon a relevant request

## Acknowledgments

We thank the Pune Knowledge Cluster members, Dr. Ajit Kembavi and Dr. Ashwini Keskar, for supporting data acquisition. We acknowledge the generous support and congratulate the PMC commissioner, officials, and staff involved in the successful COVID-19 surveillance program.

## Author contributions

VM, JM, NG, PB, SN, AJ, LS conceived the study. SN, JM, AJ performed data management and curation. DJ, PB, VM, NG conducted the data analysis, and all authors contributed to data interpretation and manuscript writing.

---

**Supplementary Table 1:** Weekly CFR in Pune, India.

| Time | Time (weeks) | Number of Patients | Number of Deaths | Cumulative number of pts | Cumulative number of deaths | Cumulative CFR (95% CI) |
| --- | --- | --- | --- | --- | --- | --- |
| **First Wave** |  |  |  |  |  |  |
| Mar-20 | 1st Week | 0 | 0 | 0 | 0 | NA |
|  | 2nd Week | 5 | 0 | 5 | 0 | NA |
|  | 3rd Week | 7 | 0 | 12 | 0 | NA |
|  | 4th Week | 25 | 4 | 37 | 4 | 10.81 (2.94 - 24.81) |
| Apr-20 | 1st Week | 112 | 24 | 149 | 28 | 19.01 (12.87 - 26) |
|  | 2nd Week | 180 | 19 | 329 | 47 | 14.29 (10.69 - 18.54) |
|  | 3rd Week | 375 | 16 | 704 | 63 | 8.95 (6.95 - 11.30) |
|  | 4th Week | 709 | 40 | 1413 | 103 | 7.29 (5.99 - 8.77) |
| May-20 | 1st Week | 618 | 50 | 2031 | 153 | 7.53 (6.42 - 8.77) |
|  | 2nd Week | 996 | 67 | 3027 | 220 | 7.27 (6.37 - 8.25) |
|  | 3rd Week | 1465 | 58 | 4492 | 278 | 6.19 (5.50 - 6.93) |
|  | 4th Week | 1961 | 87 | 6453 | 365 | 5.66 (5.10 - 6.25) |
| Jun-20 | 1st Week | 1770 | 64 | 8223 | 429 | 4.86 (4.42 - 5.33) |
|  | 2nd Week | 2115 | 83 | 10338 | 512 | 4.95 (4.54 - 5.39) |
|  | 3rd Week | 3075 | 80 | 13413 | 592 | 4.41 (4.07 - 4.77) |
|  | 4th Week | 5623 | 130 | 19036 | 722 | 3.79 (3.52 - 4.07) |
| Jul-20 | 1st Week | 6052 | 157 | 25088 | 879 | 3.50 (3.28 - 3.74) |
|  | 2nd Week | 7219 | 186 | 32307 | 1065 | 3.30 (3.10 - 3.50) |
|  | 3rd Week | 9919 | 217 | 42226 | 1282 | 3.04 (2.87 - 3.20) |
|  | 4th Week | 14142 | 355 | 56368 | 1637 | 2.90 (2.77 - 3.05) |
| Aug-20 | 1st Week | 7887 | 204 | 64255 | 1841 | 2.86 (2.74 - 3.00) |
|  | 2nd Week | 8056 | 245 | 72311 | 2086 | 2.88 (2.76 - 3.01) |
|  | 3rd Week | 8892 | 241 | 81203 | 2327 | 2.87 (2.75 - 2.98) |
|  | 4th Week | 14700 | 348 | 95903 | 2675 | 2.79 (2.69 - 2.90) |
| Sep-20 | 1st Week | 12492 | 306 | 108395 | 2981 | 2.75 (2.65 - 2.85) |
|  | 2nd Week | 14347 | 359 | 122742 | 3340 | 2.72 (2.63 - 2.81) |
|  | 3rd Week | 11762 | 300 | 134504 | 3640 | 2.71 (2.62 - 2.79) |
|  | 4th Week | 12097 | 211 | 146601 | 3851 | 2.63 (2.55 - 2.71) |
| Oct-20 | 1st Week | 5910 | 108 | 152511 | 3959 | 2.60 (2.52 - 2.68) |
|  | 2nd Week | 3997 | 78 | 156508 | 4037 | 2.58 (2.50 - 2.66) |
|  | 3rd Week | 2626 | 46 | 159134 | 4083 | 2.57 (2.49 - 2.64) |
|  | 4th Week | 3048 | 63 | 162182 | 4146 | 2.56 (2.48 - 2.63) |
| **Maintenance Phase** |  |  |  |  |  |  |
| Nov-20 | 1st Week | 1511 | 39 | 1511 | 39 | 2.58 (1.84 - 3.51) |
|  | 2nd Week | 1313 | 37 | 2824 | 76 | 2.69 (2.13 - 3.36) |
|  | 3rd Week | 2098 | 32 | 4922 | 108 | 2.19 (1.80 - 2.64) |
|  | 4th Week | 3356 | 59 | 8278 | 167 | 2.02 (1.72 - 2.34) |
| Dec-20 | 1st Week | 2169 | 48 | 10447 | 215 | 2.06 (1.79 - 2.35) |
|  | 2nd Week | 1896 | 26 | 12343 | 241 | 1.95 (1.71 - 2.21) |
|  | 3rd Week | 2014 | 58 | 14357 | 299 | 2.08 (1.85 - 2.33) |
|  | 4th Week | 2627 | 51 | 16984 | 350 | 2.06 (1.85 - 2.28) |
| Jan-21 | 1st Week | 1912 | 61 | 18896 | 411 | 2.17 (1.97 - 2.39) |
|  | 2nd Week | 1754 | 30 | 20650 | 441 | 2.13 (1.94 - 2.34) |
|  | 3rd Week | 1540 | 29 | 22190 | 470 | 2.12 (1.93 - 2.32) |
|  | 4th Week | 1823 | 66 | 24013 | 536 | 2.23 (2.05 - 2.43) |
| Feb-21 | 1st Week | 1333 | 19 | 25346 | 555 | 2.19 (2.01 - 2.38) |
|  | 2nd Week | 2171 | 35 | 27517 | 590 | 2.14 (1.98 - 2.32) |
| **Second Wave** |  |  |  |  |  |  |
| Feb-21 | 3rd Week | 2807 | 50 | 2807 | 50 | 1.78 (1.33 - 2.34) |
|  | 4th Week | 4836 | 62 | 7643 | 112 | 1.47 (1.21 - 1.76) |
| Mar-21 | 1st Week | 5970 | 54 | 13613 | 166 | 1.22 (1.04 - 1.42) |
|  | 2nd Week | 10244 | 96 | 23857 | 262 | 1.10 (0.97 - 1.24) |
|  | 3rd Week | 18479 | 180 | 42336 | 442 | 1.04 (0.95 - 1.15) |
|  | 4th Week | 35835 | 380 | 78171 | 822 | 1.05 (0.98 - 1.13) |
| Apr-21 | 1st Week | 39043 | 372 | 117214 | 1194 | 1.02 (0.96 - 1.08) |
|  | 2nd Week | 37078 | 403 | 154292 | 1597 | 1.04 (0.99 - 1.09) |
|  | 3rd Week | 37643 | 420 | 191935 | 2017 | 1.05 (1.01 - 1.10) |
|  | 4th Week | 36680 | 558 | 228615 | 2575 | 1.13 (1.08 - 1.17) |
| May-21 | 1<sup>st</sup> Week | 21021 | 304 | 249636 | 2879 | 1.15 (1.11 - 1.20) |
|  | 2<sup>nd</sup> Week | 13528 | 169 | 263164 | 3048 | 1.16 (1.12 - 1.20) |
|  | 3rd Week | 6938 | 89 | 270102 | 3137 | 1.16 (1.12 - 1.20) |
|  | 4th Week | 5391 | 47 | 275493 | 3184 | 1.16 (1.12 - 1.20) |

**Supplementary Table 2-.**
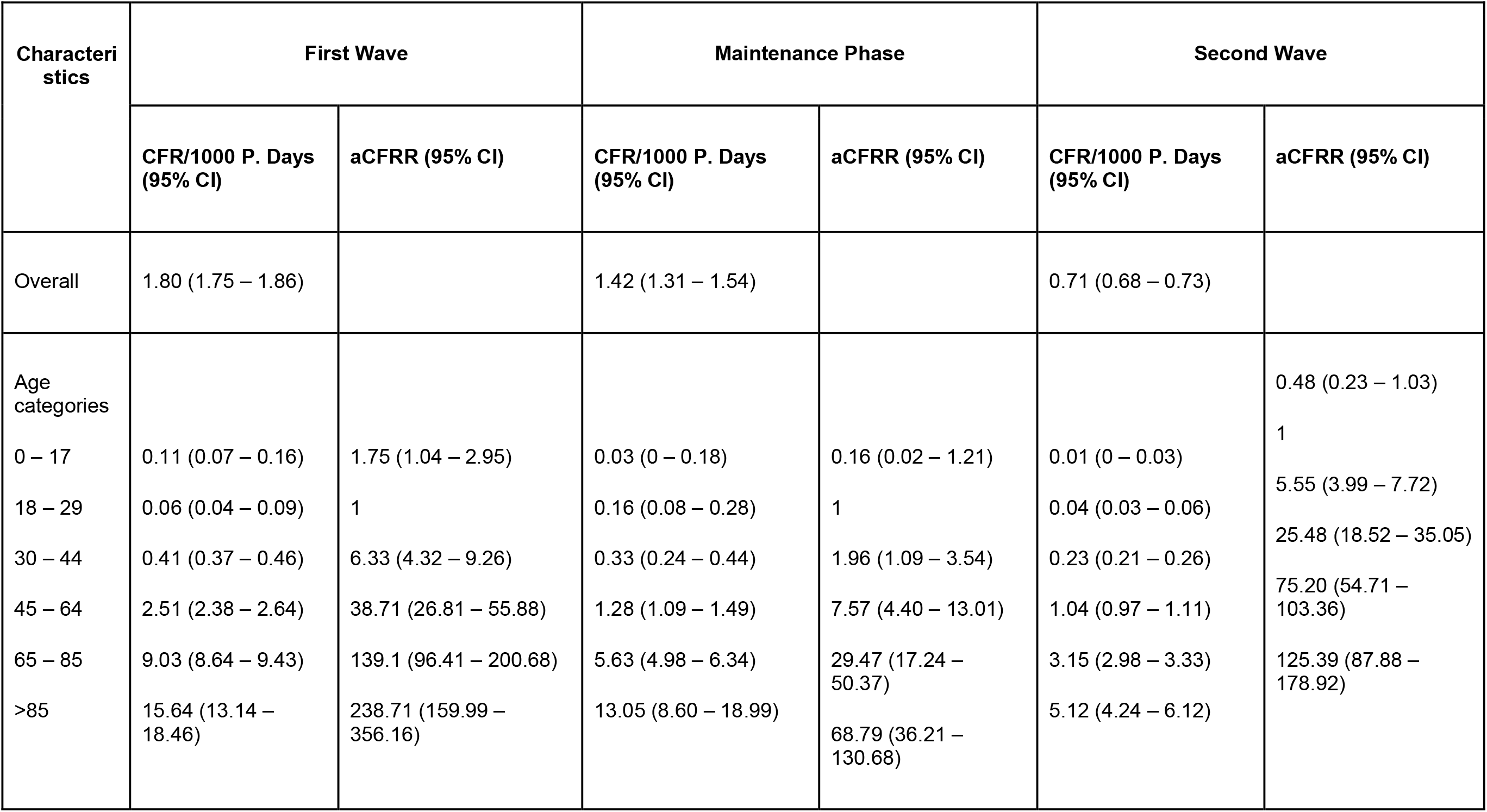
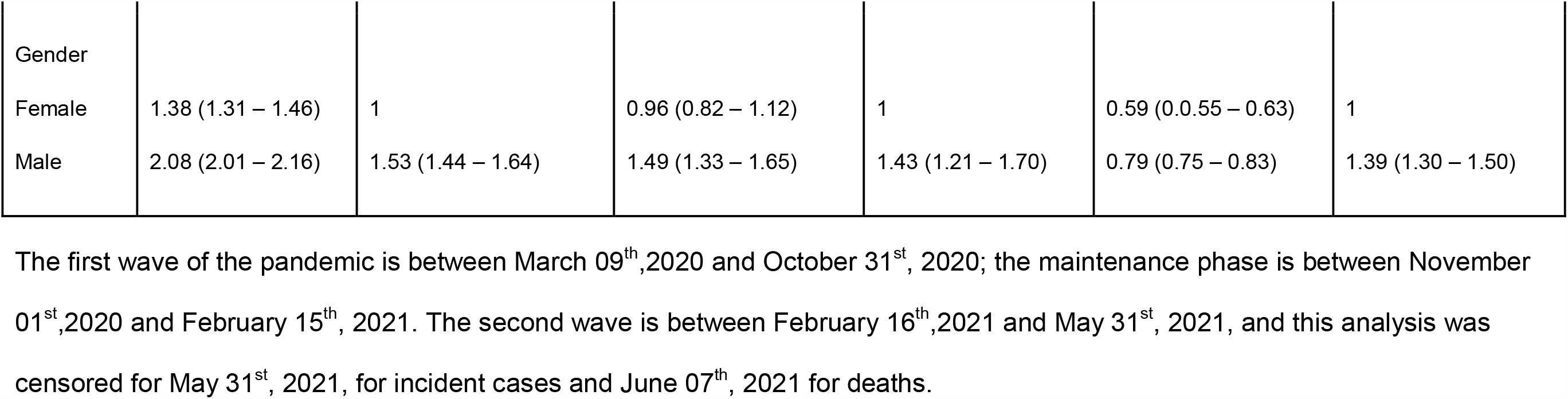
Demographic Factors Associated with COVID-19 Mortality using Poisson Regression Analysis in first wave, maintenance phase and second wave.

## SUPPLEMENTARY FIGURES

**Supplementary Figure 1-.**
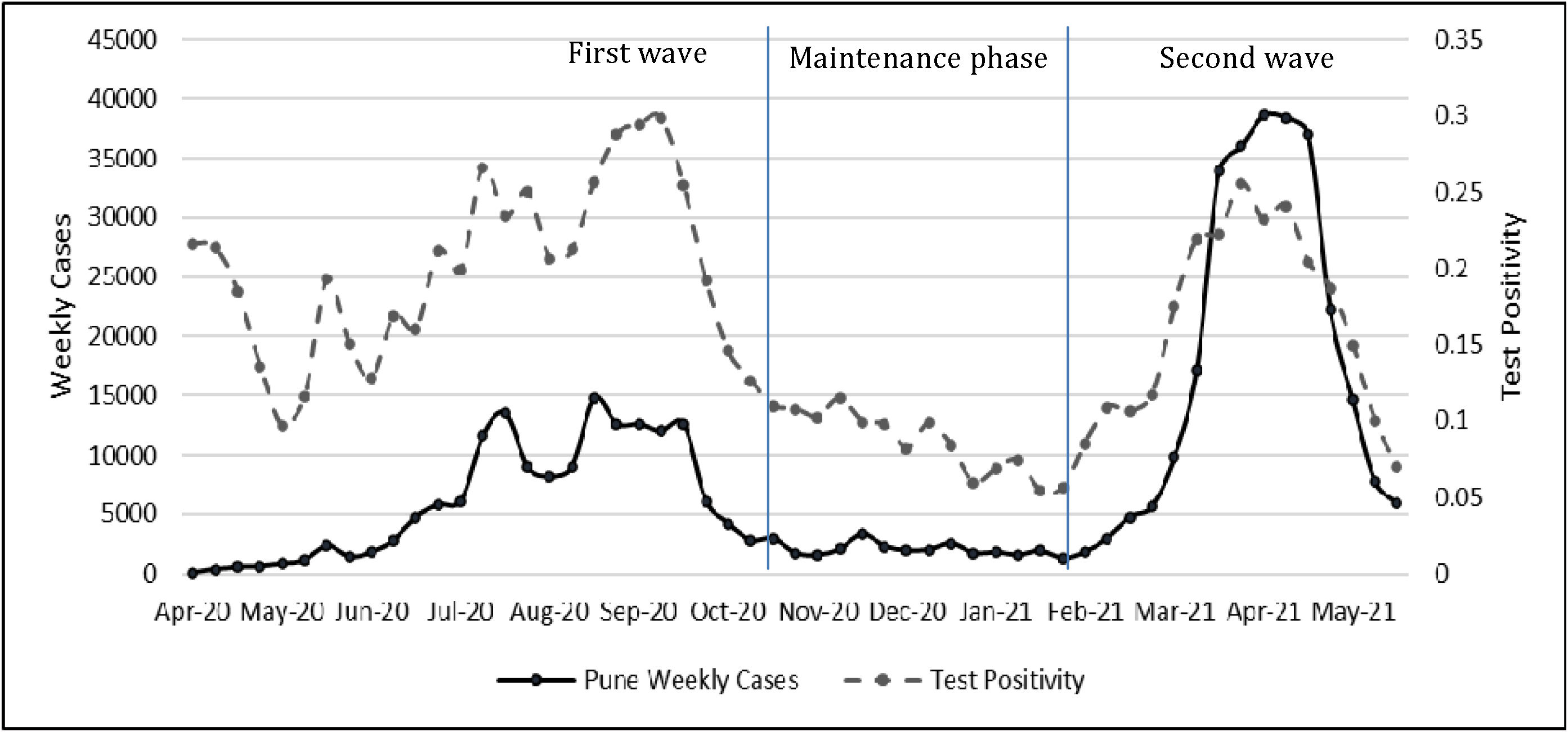
COVID-19 weekly cases and test positivity in Pune city.

**Supplementary Figure 2 –.**
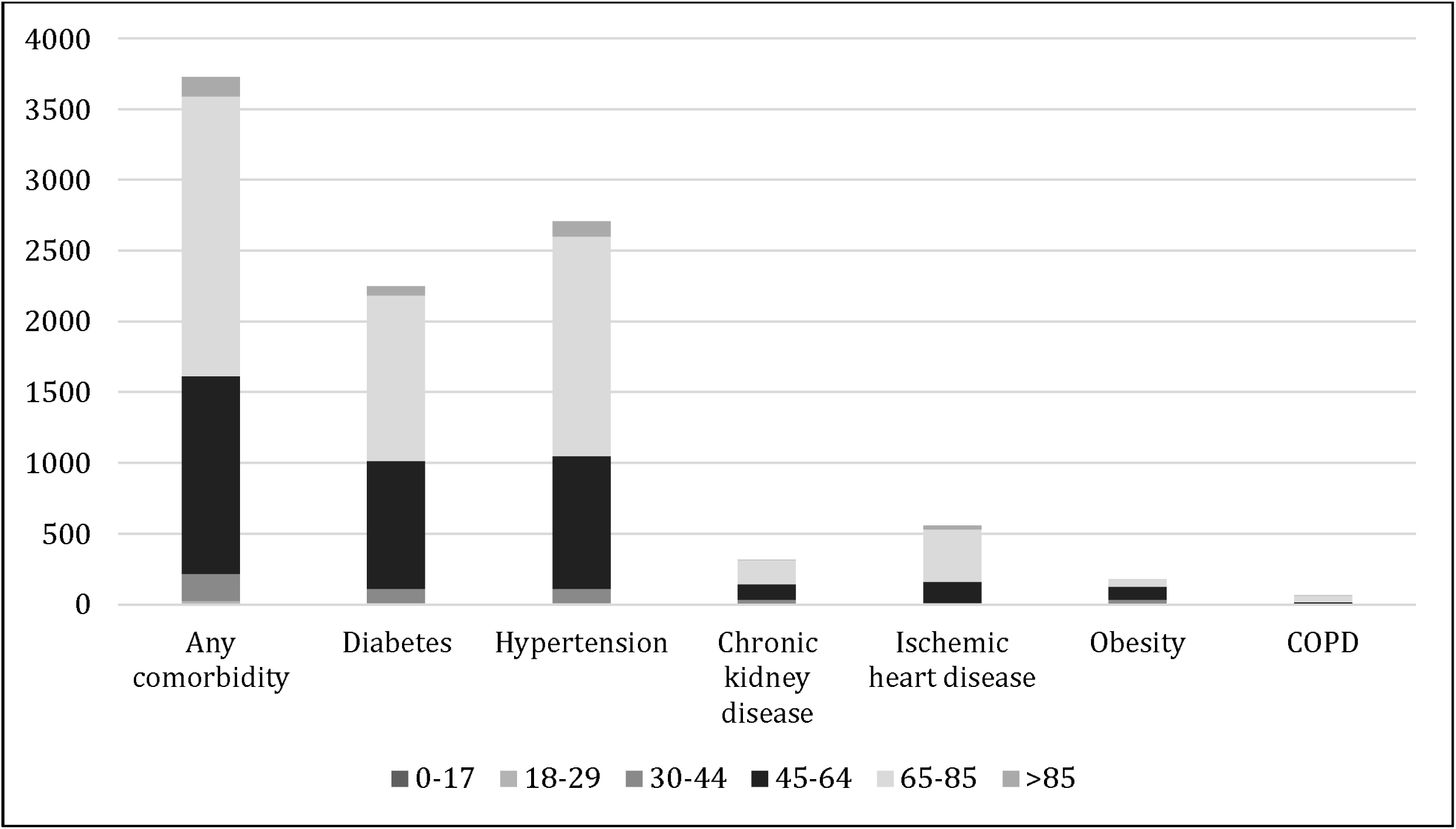
Proportion of comorbidities among COVID-19 in Pune, India – disaggregated by age groups.

